# Real-time Auditory Feedback for Improving Gait and Walking in People with Parkinson’s Disease: A Pilot and Feasibility Trial

**DOI:** 10.1101/2024.01.04.24300838

**Authors:** Nancy E. Mayo, Kedar K. V. Mate, Lesley K. Fellows, José A. Morais, Madeleine Sharp, Anne-Louise Lafontaine, Edward Ted Hill, Helen Dawes, Ahmed-Abou Sharkh

## Abstract

**Background:** Technology is poised to bridge the gap between demand for therapies to improve gait in people with Parkinson’s and available resources. A wearable sensor, Heel2Toe^TM^, a small device that attaches to the side of the shoe and gives a sound each time the person starts their step with a strong heel strike has been developed and pre-tested by a team at McGill University. The objective of this study was to estimate feasibility and efficacy potential of the Heel2Toe^TM^ sensor in changing walking capacity and gait pattern in people with Parkinson’s.

**Methods:** A pilot study was carried out involving 27 people with Parkinson’s randomized 2:1 to train with the Heel2Toe[TM] sensor and or to train with recommendations from a gait-related workbook.

**Results:** A total of 21 completed the 3-month evaluation, 14 trained with the Heel2Toe[TM] sensor and 7 trained with the workbook. Thirteen of 14 people in the Heel2Toe group improved over measurement error on the primary outcome, the Six Minute Walk Test, (mean change 66.4 m.) and 0 of the 7 in the Workbook group (mean change –19.4 m.): 4 of 14 in the Heel2Toe group made reliable change and 0 of 7 in the Workbook group. Improvements in walking distance were accompanied by improvements in gait quality. 40% of participants in the intervention group were strongly satisfied with their technology experience and an additional 37% were satisfied.

**Conclusions:** Despite some technological difficulties, feasibility and efficacy potential of the Heel2Toe sensor in improving gait in people with Parkinson’s was supported.

**Key messages regarding feasibility:** *What uncertainties existed regarding the feasibility?:* The Heel2Toe sensor had been used in clinical research as an assessment tool and in two small proof-of-concept studies with short-term supervised use to detect change and get user feedback on their experience. There was a need to test the sensor for home use and include a control group as perhaps the attention and exercise recommendations could alone have benefit. Therefore, we designed this pilot and feasibility study.

*What are the key feasibility findings?:* Dropouts from the trial were mainly related to the COVID situation. There were no adverse events in either group. Challenges with using the Heel2Toe sensor related to functionality of the app which were addressed immediately; hardware challenges were addressed in revisions including ease of charging and Bluetooth connectivity; there were challenges for people to use the smart-phone app optimally. Our current revision has removed need for the smartphone. The results also showed that people were able to use the sensor on their own at home with some technical support (average 22 minutes per person) which diminished over time and that, despite technical challenges, the majority of people were satisfied with their experience with the technology, some very much so. There was a strong response in the Heel2Toe group and a near nil response in the control group demonstrating efficacy potential.

*What are the implications of the feasibility findings for the design of the main study?:* The main study will use the revised version of the Heel2Toe sensor which has eliminated the challenges with connectivity and smartphone skills. Using the 6MWT as the outcome and based on conservative estimates of effect size (0.5), a sample size of 64 per group would be supported. This sample size would also be sufficient for estimating effects on other explanatory and downstream outcomes. Participants would keep the sensor after the study.

## Background

The disruption of the dopaminergic system in Parkinson’s Disease (PD) has a profound impact on motor networks needed to control movements.[1] Notably in people with PD, the automatic movements that typify normal walking activity are lost[2],[3] and a deteriorating a gait pattern develops characterised by quick, short, shuffling steps, narrow base of support, stooped posture, rigid trunk, and reduced arm swing. The short stride length often causes the foot to scuff the ground, causing trips and falls.[4–6] Starting, stopping, and changing direction are more difficult, gait pattern is inconsistent,[7] and freezing is common.[8] As gait impairments progress, asymmetries develop and people have difficulty adapting their walking to new or complex environments or to increased task burden.[9, 10] Walking is perceived as harder and, eventually, walking for enjoyment and health promotion abates and then ceases.

One solution to improve gait is to emphasize a heel-to-toe gait pattern,[6] something typically done during physical therapy to change posture and stride length. This strategy provides the walker with feedback and encouragement for this, usually automatic, movement. Relearning the pattern requires repeated practice and, once the therapist ceases this verbal cueing, the walker returns to their typical gait pattern.

Gait training is predominantly carried out by physical therapists with one-on-one interactions, however, there are not enough therapists for the number of people with gait vulnerabilities. Technology is poised to bridge the gap between supply and demand facilitating self-management of gait vulnerabilities. Some technologies are more successful than others, but many gaps remain in technology readiness, usability, access, training needs, and efficacy potential.

Researchers at McGill University have developed and commercialized through PhysioBiometrics Inc. a device that automates this verbal cueing by providing real-time auditory feedback when the heel strikes first when stepping. The Heel2Toe^TM^ sensor, shown in Figure 1, consists of a sensor that runs a real-time algorithm that discriminates good from poor steps with 94% accuracy,[11, 12] and generates appropriate feedback. It is classified by Health Canada as a Class I medical device (#167654). The sensor has a gyroscope, an accelerometer, and a magnetometer providing 9-degrees of freedom.

The gait cycle has been studied and described since the advent of bipedal gait.[13–15] Figure 2 presents a graphic of the normal gait cycle when tracked from the ankle joint using the gyroscope. Normal gait is characterized by two troughs and one peak. The first trough is when the ankle moves clockwise from initial contact to foot flat when there is no ankle movement allowing for weight transfer from the heel to the ball of the foot. The second trough is when the ankle again moves clockwise to push the foot off the ground to propel the body forward. Typically, the ratio of push off to heel strike is estimated at 2:1 [16, 17] The peak represents the swing phase of the gait cycle when the foot leaves the ground and swings forward to initiate another step.

The sensor detects the velocity at which the ankle moves clockwise during the initial contact of the foot during a step (angular velocity: AV). When the AV crosses a threshold for a “good step” a signal is sent via Bluetooth to a smart phone and a sound is emitted. This external positive feedback drives motor learning, retraining gait patterns to be more normal, fluid, safe, and sustainable. To normalize walking, people must relearn motor sequences and develop needed adjuncts to efficient walking: strength, power, core stability, balance, etc. Physical therapy targets adjuncts but motor learning requires instruction, repetition, and practice.[18] At least some of the neural mechanisms underlying this learning are likely aberrant in PD. Motor learning via feedback involves neuroplasticity in corticostriatal and striato-cerebellar circuits in a partially dopamine-dependant manner.[18–21]

In two proof-of-concept studies of 6 people with PD[22] and 6 pre-frail seniors[23] receiving 5 training sessions with Heel2Toe^TM^ over 2 weeks, every person made at least one clinically meaningful change on one gait parameter after training. The potential mechanism of action is a dopamine-driven reward and feedback loop.[24] Here we set out to estimate the extent to which training with the Heel2Toe^TM^ over a longer period of time (3 months) was feasible and acceptable to participants and to estimate changes in walking capacity and gait pattern among people training with feedback from the sensor and among those training without feedback.

The hypotheses for which the pilot trial will provide supporting data is that people in the group training with feedback from the Heel2Toe sensor will make greater gains in walking capacity and motivation and will show more optimal changes in parameters of gait quality than will be observed in the control group.

## Design

A two-group, 2:1 randomized, feasibility trial was carried out with repeated measures of gait parameters and walking outcomes. The randomization sequence was generated by an independent statistician. The trial was prospectively registered on April 3, 2020 under the name “Improving Walking With Heel-To-Toe Device” on ClinicalTrials.gov (NCT04300348) https://register.clinicaltrials.gov/prs/app/action/SelectProtocol?sid=S0009NRV&selectaction=Edit&uid=U0000572&ts=2&cx=-nba3sj; The project was approved by the Research Ethics Board of the McGill University Health Center.

The feasibility phase followed the recommendations from the CONSORT extension to randomised pilot and feasibility trials (PAFS).[25, 26] PAFS emphasizes testing all aspects of data collection and processes of the intervention and measurement, but warns against between-group testing of efficacy due to lack of statistical power.

## Population

People with PD manifesting gait impairments and meeting the criterion that usual walking is without a walking aid[27], corresponding to Hoehn and Yahr Scale of 2 to 3, were recruited from the Movement Disorders Clinics at McGill sites and the Quebec Parkinson Network. Patients with documented cognitive impairment based on their recorded score on the Montreal Cognitive Assessment (MOCA)[28] were not approached for inclusion. All patients kept their usual dopaminergic medication schedule throughout the study. People were assessed at a time that corresponded to their medication regimen.

## Intervention

Both groups received a workbook with instructions on simple exercises to facilitate a better walking pattern (available at physiobiometrics.com), 5 sessions with a physiotherapist (PT) over two weeks to practice walking well and four specific exercises, one for each major joint area involved in walking (foot and ankle, knees, hip, trunk). This personal gait training period was followed by independent home practice over 3 months. Both groups were instructed to practice walking with the sensor for a minimum period of 5 minutes, twice a day. The exercises were to be done before each walk, 10 to 15 repetitions, During the 5 therapy sessions, the Heel2Toe group was taught to trigger the sensor with a strong heel strike to receive the feedback and how to use the sensor and the app on the smartphone. This instruction was in preparation for independent home use for 3 months. The Workbook group also received similar verbal instruction during these 5 therapy sessions when walking with the Heel2Toe sensor but received no feedback from the sensor.

## Measures

The primary outcome was the 6 Minute Walk Test (6MWT), a performance-based outcome (PerfO) of functional walking capacity.[29] A secondary PerfO was the Standardized Walking Obstacle Course (SWOC)[30], a timed performance-based test involving starting, stopping, turning, and making motor decisions. Average values for people with mean age 63 years is reported to be 12 seconds [31] Sit-to-Stand, the number completed in 30 seconds, was also assessed. The average for people aged 70-74 years is reported to range for 10 to 17 depending on sex.[32] Assessors were unaware of the group assignment at time of assessment.

Data on constructs related to other aspects of brain health (motivation, symptoms, function and quality of life) were also collected using patient-reported outcome measures (PROMS), Motivation was measured using the Starkstein Apathy Scale [33] and an inventory of activities based on the World Health Organizations International Classification of Functioning, Disability and Health (ICF). From this ICF bank of 393 activity and participation items, 17 were chosen as relevant for this context and rated based on degree of self-initiation (0 to 2) and degree of effort (0-2). This measure is under development and this study provides feasibility data to support further directions.

Symptoms of anxiety, depression, pain, and energy and quality of life were measured using Visual Analogue Health States[34] on a 0 to 100 scale with higher values indicating better health states. Values less than 60 would be considered to reflect a clinical situation where treatment might be indicated, equivalent to a value of >40 when the scale is reversed [35, 36]. Function was measured with the NeuroQOL[37]; health related quality of life (HRQL) was measured with the 8-item Parkinson Deficit Questionnaire (PDQ)[38] where higher scores indicate poorer HRQL, and the EuroQol measure.[39]

Indicators of gait quality were obtained directly from the Heel2Toe sensor during the 6MWT. Due to inconsistent Bluetooth connection from sensor to smart phone (fixed over the course of the trial), the number of recorded steps varies. The indicators are: percentage good steps (those that passed the pre-determined threshold of –150°/sec of ankle angular velocity (AV); AV at each part of the gait cycle (heel strike, push off, swing) and associated coefficients of variation (CV) of AV, where CV is calculated as the ratio of standard deviation (SD) to the mean of each parameter, expressed as a percentage. Two measures were derived from these data: power phase, the area of the two troughs under the under the zero AV line (ankle still during stance) termed area under the line (AUL); and balance phase, the area above the zero-line termed area above the curve (AAL). The balance phase is so named as its shape and area is determined by the ability of the person to do single-leg stance long enough and lift the swing leg high enough for the foot to clear the ground. Average time in swing and CV were also measured. A total of 13 gait quality parameters are reported here.

To identify whether gait quality parameters changed over the intervention period, a difference of 10% from baseline to 3 months was used as the critical value. A 10% change from baseline indicates important change in different types of measures.[40, 41] including gait parameters.[42]

## Analysis

Reliable change[43], magnitude of change relative to pre-post variability and observed inter-test correlation, was calculated for each participant within each group, over the intervention and follow-up periods. The critical value for a single arm, pre-post, study is 1.645. Also presented are results from a paired-t test and effect sizes[44] for each group. The sample was too small to use imputation as needed for an intention-to-treat analysis and so only per protocol, within group, results are presented. Data on secondary outcomes are presented for descriptive purposes only as sample sizes are small, and variability large. As this was a pilot study, no between group analyses are indicataed[25, 26] but estimates of change were used to guide power for a future trial.

Data on gait quality parameters are presented per person according to group. The number of gait parameters showing improvement, no change, or deterioration were summed for each person and accumulated over all people. Rate of improvement per group was calculated as total number of improved gait parameters divided by the total number of person-measures assessed (parameters*people); 95% confidence intervals (CI) for these rates were calculated.

## Sample size

The study was powered to detect a minimal important within-group change of moderate or greater magnitude (effect size ½ standard deviation) on the 6MWT. A sample size of 20 per group was targeted to provide 80% power (Type I error 0.05) to provide 95% confidence that future estimates of within-group effect will exclude the null value of 0 correlation (95% CI: 0.03 to 0.96 SD). The trial was approved to start on the day that McGill University shut down because of COVID (March 2020). The trial was not permitted to start with in-person assessments and therapy sessions until April 2021 and funding restrictions required the trial duration to be curtailed resulting in a reduced sample size. Thus, we chose to assign people 2:1 to the intervention and control groups to maximise the number receiving intervention.

## Results

Figure 3 shows the path of participants through the study. Of the 33 eligible, 27 agreed. As the study was curtailed because of COVID, we did not attempt to recruit others. Of these, 18 were randomized to the Heel2Toe group and 9 to the Workbook group. One person in the Heel2Toe group did not receive any intervention owing to difficulty with scheduling. Fourteen people in the Heel2Toe group completed the 3-month assessment and 13 completed the 6-month assessment. In the Workbook group, these numbers were 7 and 6. Reasons for incompletion related to the demands of the trial, fear of COVID, travel, and illness.

The characteristics of the participants in terms of demographics and brain health outcomes at randomization are shown in Table 1. There were some qualitative differences on symptoms such as pain, fatigue, and mood with the participants in the Workbook group reporting average values in the range of clinical concern; however, there was a considerable amount of variability in the ratings.

**Table 1:**
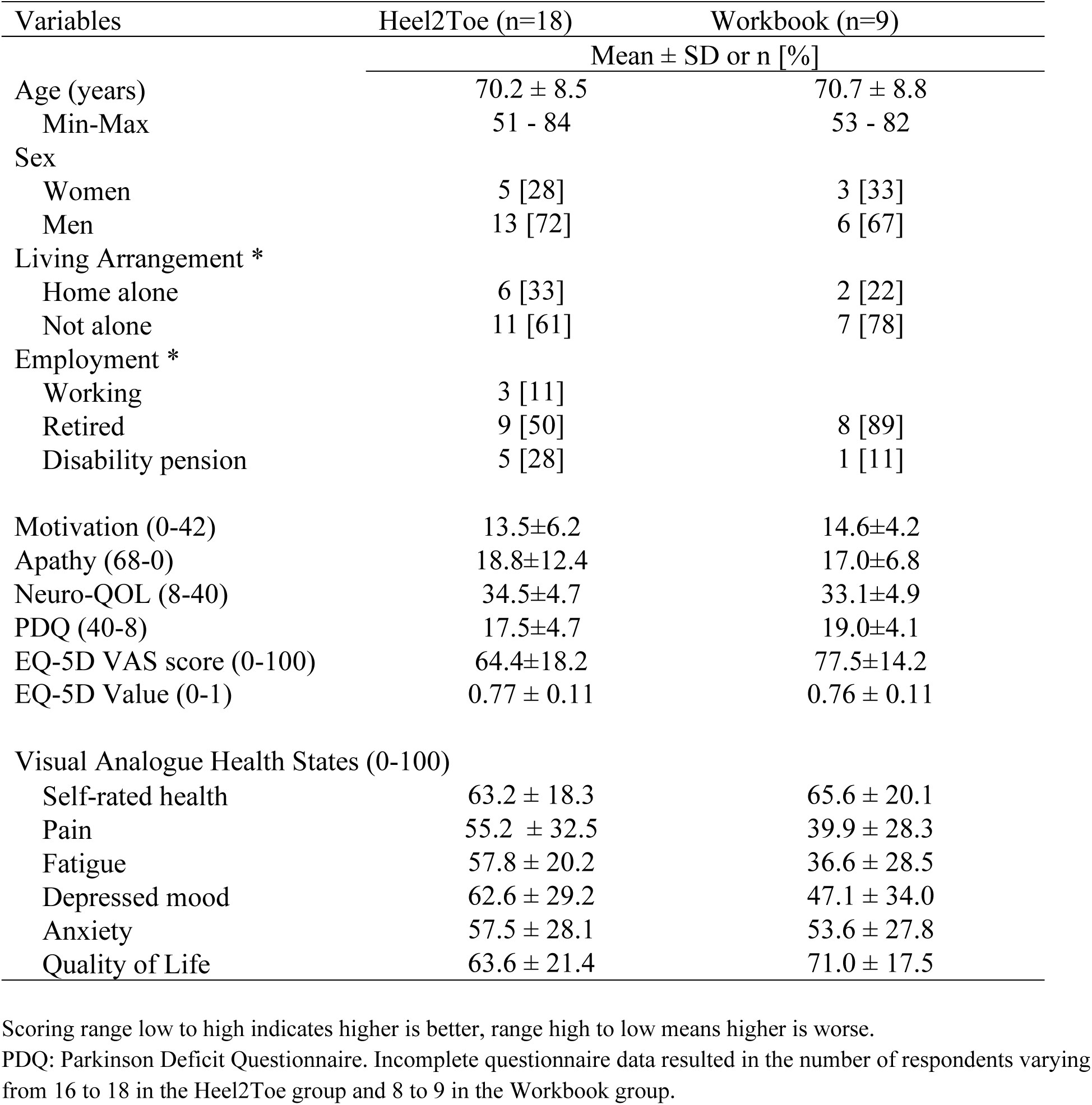
Characteristics of Participants in Each of the Two Groups at Randomization.

| Variables | Heel2Toe (n=18) | Workbook (n=9) |
| --- | --- | --- |
| | Mean ± SD or n [%] | |
| Age (years) | 70.2 ± 8.5 | 70.7 ± 8.8 |
| Min-Max | 51 - 84 | 53 - 82 |
| Sex |  |  |
| Women | 5 [28] | 3 [33] |
| Men | 13 [72] | 6 [67] |
| Living Arrangement * |  |  |
| Home alone | 6 [33] | 2 [22] |
| Not alone | 11 [61] | 7 [78] |
| Employment * |  |  |
| Working | 3 [11] |  |
| Retired | 9 [50] | 8 [89] |
| Disability pension | 5 [28] | 1 [11] |
| Motivation (0-42) | 13.5±6.2 | 14.6±4.2 |
| Apathy (68-0) | 18.8±12.4 | 17.0±6.8 |
| Neuro-QOL (8-40) | 34.5±4.7 | 33.1±4.9 |
| PDQ (40-8) | 17.5±4.7 | 19.0±4.1 |
| EQ-5D VAS score (0-100) | 64.4±18.2 | 77.5±14.2 |
| EQ-5D Value (0-1) | 0.77 ± 0.11 | 0.76 ± 0.11 |
| Visual Analogue Health States (0-100) |  |  |
| Self-rated health | 63.2 ± 18.3 | 65.6 ± 20.1 |
| Pain | 55.2 ± 32.5 | 39.9 ± 28.3 |
| Fatigue | 57.8 ± 20.2 | 36.6 ± 28.5 |
| Depressed mood | 62.6 ± 29.2 | 47.1 ± 34.0 |
| Anxiety | 57.5 ± 28.1 | 53.6 ± 27.8 |
| Quality of Life | 63.6 ± 21.4 | 71.0 ± 17.5 |

Scoring range low to high indicates higher is better, range high to low means higher is worse.
PDQ: Parkinson Deficit Questionnaire. Incomplete questionnaire data resulted in the number of respondents varying from 16 to 18 in the Heel2Toe group and 8 to 9 in the Workbook group.

The results on the primary outcome, the 6MWT, are presented for each group separately in Table 2. Average values at baseline were approximately 75-80% of what would be predicted for age. The number of participants differs at each timepoint because of missing data. Among the 14 people in the Heel2Toe group with both baseline and three-month evaluations, the average change in the 6MWT was 66.4 meters (SD: (55.6); the change 6MWT for the 7 people in the Workbook group was –19.4 meters (SD: 41.6). The difference in the Heel2Toe group was associated with a paired t-test value of +4.47 (p=0.0006) and an effect size of +0.47; the corresponding effect parameters for the Workbook group were –1.24 (p=0,26) and –0.11, respectively. These parameters are also presented at the 6-month follow-up visit. The proportion of people in the Heel2Toe group who improved more than measurement error for was 13/14 after 3 months and 12/13 after 6 months; for the Workbook group, these ratios were 0/7 and 5/6. The average change at follow-up in the Heel2Toe group was 75.7 m (SD: 81), and the average change in the Workbook group was 34.4 m (95.7 m). However, reliable change was 4/14 and 5/13 for these two time periods for the Heel2Toe group and 0/7 and 1/6, for the Workbook group. Individual changes to 3 and to 6 months on the 6MWT are presented in Supplementary Figures 1a and 1b for each of the two groups. In the Heel2Toe group, most of the changes observed over the active intervention period were maintained to 6 months. For the Workbook group, one person made a dramatic change, resulting in a large mean change, others made some smaller changes.

**Table 2.** Results on the 6MWT and other Performance Measures for Each Group.

| Variables | Heel2Toe (n=18)<br>Mean ± SD [n] | Workbook (n=9)<br>Mean ± SD [n] |
| --- | --- | --- |
| **6MWT<sup>1</sup> [Average for age 70]** |  |  |
| Baseline [450-540 m.] | 381.0±140.1 [16] | 409.3±183.4 [9] |
| Post-intervention (3 months) | 451.0±127.7 [14] | 370.1±198.5 [7] |
| Change from baseline | 66.4±55.6 [14] | -19.4±41.6 [6] |
| Paired t-test | +4.47 (p=0.0006) | -1.24 (p=0.26) [6] |
| Effect Size | +0.47 | -0.11 |
| Follow-up (6 months) | 475.8±83.2 [10] | 425.7±200.1 [6] |
| Change from baseline | 75.7±81.1 [10] | 34.4±95.7 [6] |
| Paired t-test | +2.95 (p=0.016) | -0.88 (p=0.42) [6] |
| Effect Size | +0.54 | +0.19 |
| Change >20 m. n (%) |  |  |
| Baseline to Post-intervention | 13/14 | 0/7 |
| Baseline to Follow-up | 12/13 | 5/6 |
| Reliable change: n (%) |  |  |
| Baseline to Post-intervention | 4/14 | 0/7 |
| Baseline to Follow-up | 5/13 | 1/6 |
| **Time (seconds) to complete first SWOC<sup>2</sup> [average for healthy]** |  |  |
| Baseline [12 seconds] | 19.6±12.12 [19] | 29.5±14.6 [9] |
| 3 months | 23.6±10.0 [15] | 27.0±11.0 [7] |
| Change | 4.3±7.1 [15] | -6.1±8.3 [7] |
| 6 months | 19.9±6.1 [14] | 25.5±10.8 [7] |
| **N of SWOC<sup>8</sup> Trials Completed at Baseline** |  |  |
| 1 / 2 / 3 | 1 / 2 / 6 | 0 / 2 / 4 |
| 4 / 5 / 6 | 4 / 3 / 2 | 1 / 1 / 1 |
| Increase trials at 3 months | 5/15 (33.3%) | 1/7 (14.3%) |
| **Sit-to-Stand<sup>3</sup> (n in 30 seconds) [Average for age 70]** |  |  |
| Baseline [10-17] | 11.4±4.9 [18] | 10.4±4.8 [9] |
| 3 months | 13.0±6.1 [14] | 10.0±7.2 [7] |
| Change | 2.1±5.2 [14] | 0.14±3.3 [7] |
| 6 months | 13.8±5.2 [13] | 12.2±4.2 [6] |

Effect Size: change / SD at baseline. Numbers in square brackets are the numbers of people with data on the measure at each time point [n]. Estimates from https://www.omnicalculator.com/health/6-minute-walk-test for age 70 man and woman of average height and weight. 2. Standardized Walking Obstacle Course measured in seconds with higher worse; people also asked if they desired to repeat the course to get a better time. 3. Number in 30 seconds; more is better, average is 10-17 for age 70-74 years both sexes combined.

Table 2 also presents the results on the other PerfOs, the SWOC and Sit-to-Stand. The time to complete the obstacle course was on average 20 to 30 seconds across groups with normal values reported as 12 seconds. Of interest is that the number of people agreeing to make more attempts on this course was greater after the intervention with the Heel2Toe sensor (33%) whereas only 14.3% chose this option in the Workbook group. Results on the Sit-to-Stand test were at the lower end of normal for age.

Values on PROMs for motivation and other brain health outcomes are presented in Table 3. Observed change on the Apathy Inventory, improvement in the Heel2Toe group (–4.2±7.6) and worsening in the Workbook group (3.6±10.9) supported our hypothesis that the feedback affects motivation. Also, overall rating of health and health-related quality of life as measured by the EQ-5D VAS and EQ-5D utility improved in the Heel2Toe group (VAS: +7.1±10.1; utility: 0.01±0.8) and worsened in the Workbook group VAS: –10.2±12.7; utility: –0.8±0.12). Other brain health outcomes that changed in hypothesized directions were anxiety, pain, and overall quality of life. There were no adverse events in either group.

**Table 3.**
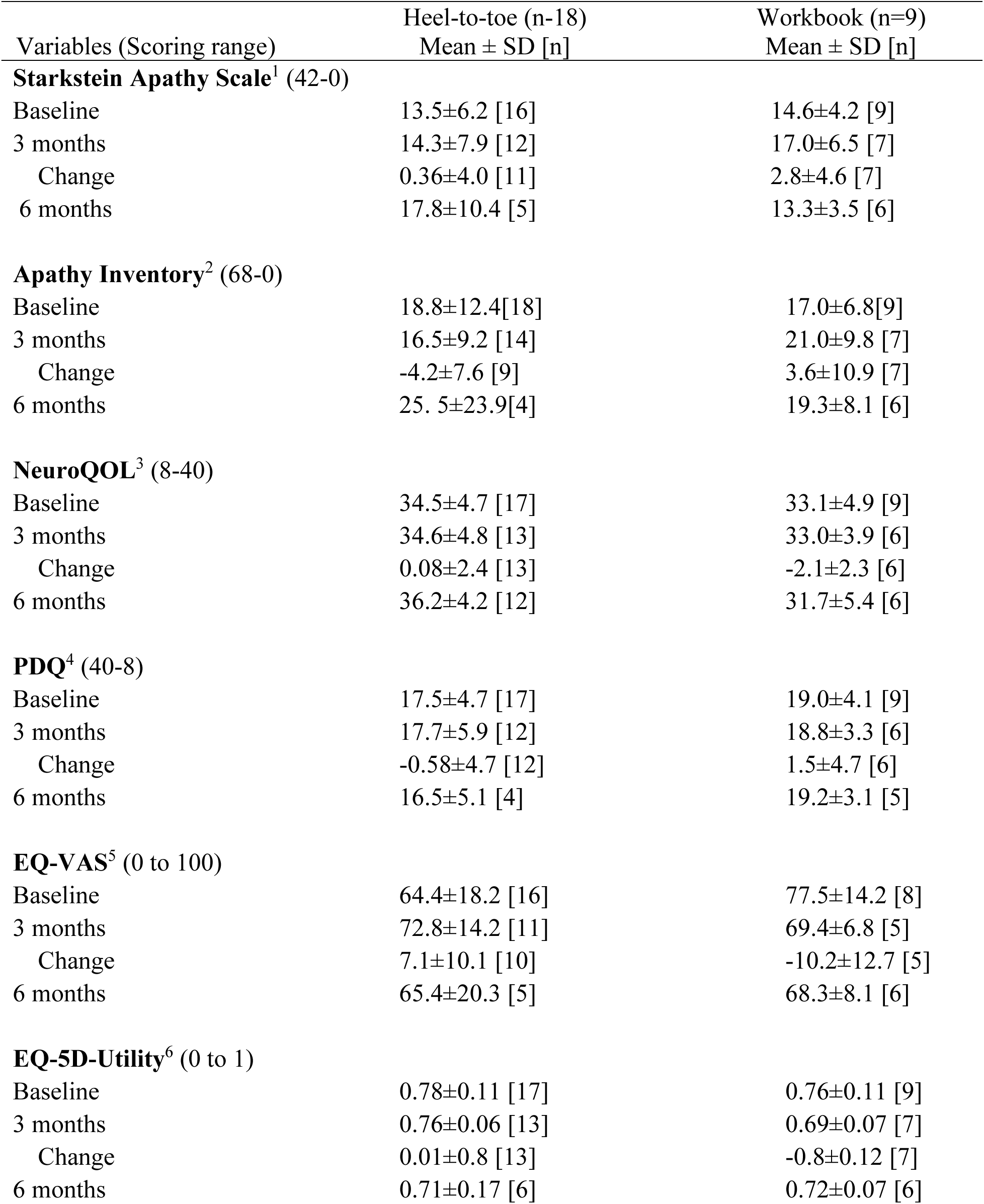
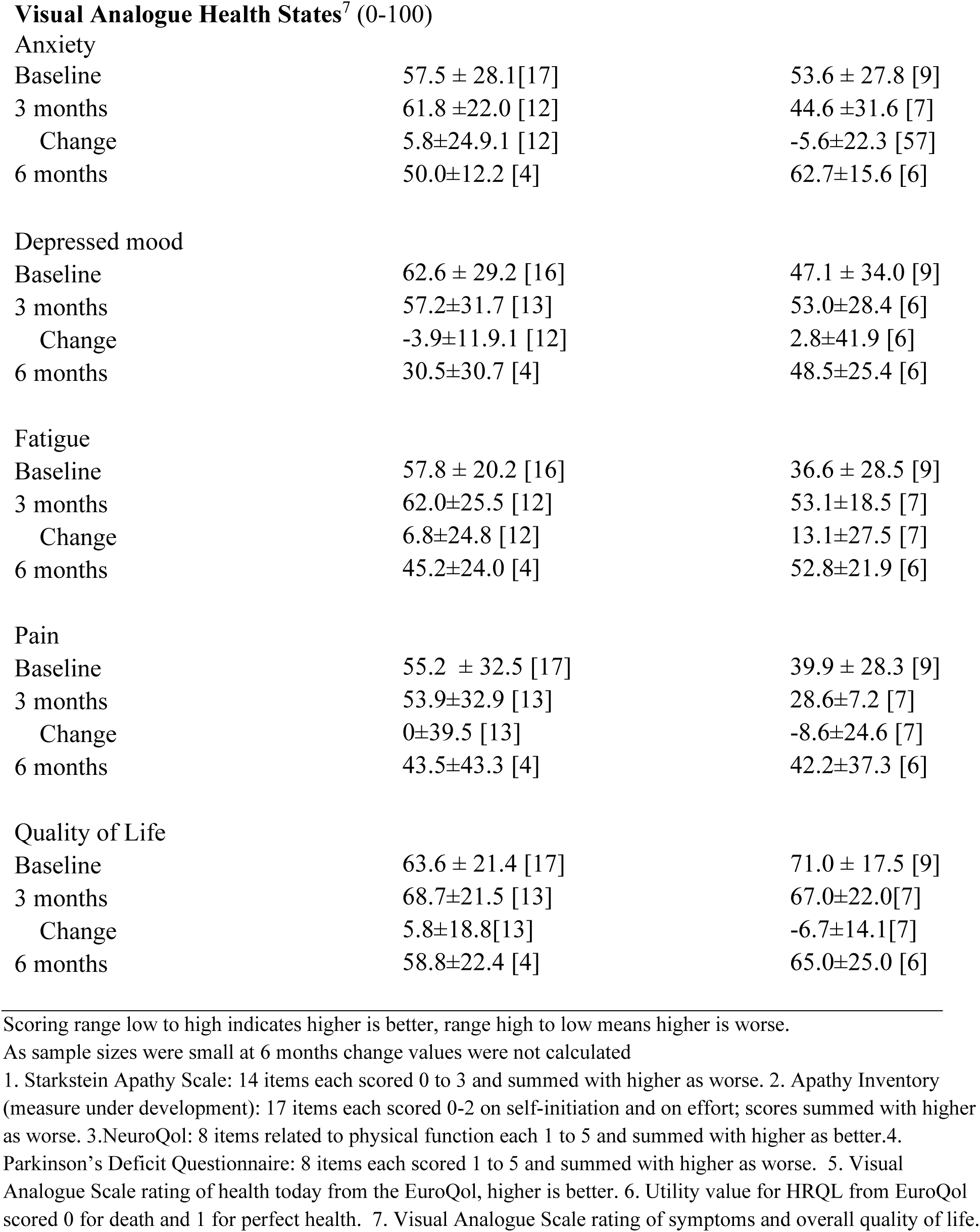
Brain Health Outcomes for Participants in Each Group Over Time.

| Variables (Scoring range) | Heel-to-toe (n=18)<br>Mean ± SD [n] | Workbook (n=9)<br>Mean ± SD [n] |
| --- | --- | --- |
| **Starkstein Apathy Scale<sup>1</sup> (42-0)** |  |  |
| Baseline | 13.5±6.2 [16] | 14.6±4.2 [9] |
| 3 months | 14.3±7.9 [12] | 17.0±6.5 [7] |
| Change | 0.36±4.0 [11] | 2.8±4.6 [7] |
| 6 months | 17.8±10.4 [5] | 13.3±3.5 [6] |
| **Apathy Inventory<sup>2</sup> (68-0)** |  |  |
| Baseline | 18.8±12.4[18] | 17.0±6.8[9] |
| 3 months | 16.5±9.2 [14] | 21.0±9.8 [7] |
| Change | -4.2±7.6 [9] | 3.6±10.9 [7] |
| 6 months | 25.5±23.9[4] | 19.3±8.1 [6] |
| **NeuroQOL<sup>3</sup> (8-40)** |  |  |
| Baseline | 34.5±4.7 [17] | 33.1±4.9 [9] |
| 3 months | 34.6±4.8 [13] | 33.0±3.9 [6] |
| Change | 0.08±2.4 [13] | -2.1±2.3 [6] |
| 6 months | 36.2±4.2 [12] | 31.7±5.4 [6] |
| **PDQ<sup>4</sup> (40-8)** |  |  |
| Baseline | 17.5±4.7 [17] | 19.0±4.1 [9] |
| 3 months | 17.7±5.9 [12] | 18.8±3.3 [6] |
| Change | -0.58±4.7 [12] | 1.5±4.7 [6] |
| 6 months | 16.5±5.1 [4] | 19.2±3.1 [5] |
| **EQ-VAS<sup>5</sup> (0 to 100)** |  |  |
| Baseline | 64.4±18.2 [16] | 77.5±14.2 [8] |
| 3 months | 72.8±14.2 [11] | 69.4±6.8 [5] |
| Change | 7.1±10.1 [10] | -10.2±12.7 [5] |
| 6 months | 65.4±20.3 [5] | 68.3±8.1 [6] |
| **EQ-5D-Utility<sup>6</sup> (0 to 1)** |  |  |
| Baseline | 0.78±0.11 [17] | 0.76±0.11 [9] |
| 3 months | 0.76±0.06 [13] | 0.69±0.07 [7] |
| Change | 0.01±0.8 [13] | -0.8±0.12 [7] |
| 6 months | 0.71±0.17 [6] | 0.72±0.07 [6] |
| **Visual Analogue Health States<sup>7</sup> (0-100)** |  |  |
| Anxiety |  |  |
| Baseline | 57.5 ± 28.1 [17] | 53.6 ± 27.8 [9] |
| 3 months | 61.8 ± 22.0 [12] | 44.6 ± 31.6 [7] |
| Change | 5.8 ± 24.9 [12] | -5.6 ± 22.3 [57] |
| 6 months | 50.0 ± 12.2 [4] | 62.7 ± 15.6 [6] |
| Depressed mood |  |  |
| Baseline | 62.6 ± 29.2 [16] | 47.1 ± 34.0 [9] |
| 3 months | 57.2 ± 31.7 [13] | 53.0 ± 28.4 [6] |
| Change | -3.9 ± 11.9 [12] | 2.8 ± 41.9 [6] |
| 6 months | 30.5 ± 30.7 [4] | 48.5 ± 25.4 [6] |
| Fatigue |  |  |
| Baseline | 57.8 ± 20.2 [16] | 36.6 ± 28.5 [9] |
| 3 months | 62.0 ± 25.5 [12] | 53.1 ± 18.5 [7] |
| Change | 6.8 ± 24.8 [12] | 13.1 ± 27.5 [7] |
| 6 months | 45.2 ± 24.0 [4] | 52.8 ± 21.9 [6] |
| Pain |  |  |
| Baseline | 55.2 ± 32.5 [17] | 39.9 ± 28.3 [9] |
| 3 months | 53.9 ± 32.9 [13] | 28.6 ± 7.2 [7] |
| Change | 0 ± 39.5 [13] | -8.6 ± 24.6 [7] |
| 6 months | 43.5 ± 43.3 [4] | 42.2 ± 37.3 [6] |
| Quality of Life |  |  |
| Baseline | 63.6 ± 21.4 [17] | 71.0 ± 17.5 [9] |
| 3 months | 68.7 ± 21.5 [13] | 67.0 ± 22.0 [7] |
| Change | 5.8 ± 18.8 [13] | -6.7 ± 14.1 [7] |
| 6 months | 58.8 ± 22.4 [4] | 65.0 ± 25.0 [6] |

Scoring range low to high indicates higher is better, range high to low means higher is worse.
As sample sizes were small at 6 months change values were not calculated
1. Starkstein Apathy Scale: 14 items each scored 0 to 3 and summed with higher as worse. 2. Apathy Inventory (measure under development): 17 items each scored 0-2 on self-initiation and on effort; scores summed with higher as worse. 3. NeuroQol: 8 items related to physical function each 1 to 5 and summed with higher as better. 4. Parkinson's Deficit Questionnaire: 8 items each scored 1 to 5 and summed with higher as worse. 5. Visual Analogue Scale rating of health today from the EuroQol, higher is better. 6. Utility value for HRQL from EuroQol scored 0 for death and 1 for perfect health. 7. Visual Analogue Scale rating of symptoms and overall quality of life.

Supplemental Tables 1 and 2 present the results of the analyses on the gait quality parameters. Despite small numbers, the groups were relatively well balanced at baseline (ST1). Values on gait quality parameters are presented at baseline and at the end of the active intervention period (3 months) for each person according to group (ST2). Values that differed by 10% were coloured coded green for improvement, yellow for no change, and orange for deterioration. Supplemental Figure 1 shows the proportion of participants in the two groups who, over 3 months active intervention, improved, remained the same, or deteriorated on gait parameters. The rate of improvement in the Heel2Toe group was 49.7% (95% CI: 39.6% to 61.5%) and 13.5% in the Workbook group (95% CI: 5.4% to 27.7%).

The results on usability of the Heel2Toe sensor are also presented in Supplementary Figure 2 (data on use) and Supplementary Figure 3 (satisfaction).

## Discussion

This study found strong evidence that that walking training with feedback from the Heel2Toe[TM] sensor is a feasible intervention. The results, shown in Table 2 and Supplementary Figure 1a, strongly support efficacy potential based for the Heel2Toe sensor based on the magnitude of average change in each of the two groups (+66.4 vs. –19.4), the proportion of people making change greater than measurement error (13/14 vs. 0/7) and in proportion making reliable change (4/14 vs. 0/14). The inclusion criteria for this pilot were broad and we found important improvements across the range of baseline walking capacity.

Information on usability pointed out areas for revision of the Heel2Toe[TM] sensor all of which have now been implemented into the latest version. The results also showed that people were able to use the sensor on their own at home with some technical support which diminished over time and that, despite technical challenges, the majority of people were satisfied with their experience with the technology. We also found that there were some changes in motivation favouring the Hee2Toe group which is considered to be one of the mechanisms contributing to improved outcomes.

There is support from the literature for the effectiveness of biofeedback in improving gait patterns in healthy and clinical populations including people with PD[45, 46] but few feedback devices are available to the general public.[47]

This study was designed to provide evidence as to feasibility and hence its limitations relate specifically to that design. Large effect in the intervention group, notwithstanding, the sample size was small particularly in the control group and the study was not powered for between-group comparisons. The planned sample size was not achieved because of the multiple delays and protocol changes owing to the volatile COVID situation during the study period. This also affected retention into the trial as people were worried about contagion and had other stressful situations to deal with.

The number of technical issues uncovered was both a limitation and a strength as we were able to modify some in real-time and others for subsequent revision.

We used a measure of motivation (apathy) that is under development to obtain data on its performance in this population. Results from this early deployment should be interpreted with caution.

Based on the results, the feasibility of a pragmatic definitive trial is supported. Using the 6MWT as the outcome and based on conservative estimates of effect size (0.5), a sample size of 64 per group would be supported. This sample size would also be sufficient for estimating effects on other explanatory and downstream outcomes.

List of Figures

Figure 1: Heel2Toe[TM] sensor

Figure 2. Typical Gait Cycle

Figure 3. Flow of Participants through the Study

## Supplementary Appendix

Indicators of Usability

The total number of calls among the 18 participants in the Heel2toe group over the whole intervention period was 117, ranging from 1 to 29 calls, averaging 7 per person and the total time was 344 minutes, ranging from 2 to 70 minutes for an average of 22 per person. Each person experienced some challenges with the sensor which were corrected either immediately or for the next participant. Challenges related to functionality of the app were addressed immediately; hardware challenges were addressed in revisions including ease of charging, Bluetooth connectivity, and attachment.

Supplementary Figure 2 presents the distribution of Heel2Toe use for each participant in the intervention group over the 2-week training period (blue bars) and over the 3-month home practice (orange bars). All participants were instructed to use the Heel2Toe sensor daily. The proportion of days of use during the training period ranged from 0% (person A) to >180% (person P). The median usage value was 54%, located between person H and I. The orange bars show the usage for the period of home practice. Usage in the training period was not carried over into the home training period: the range was from 0% to >100% and the median value was 37%, located at person M.

Supplementary Figure 3 shows that, while there were technical challenges with the sensor and there was a learning curve, the majority of people were satisfied and would recommend it to others.

## Declarations

### Ethics approval ad consent to participate

The project was approved by the Research Ethics Board of the McGill University Health Center on Feb 17, 020 (File # 2020-5842).

## Consent for publication

All authors have reviewed and approved the manuscript.

## Availability of data and material

Data could be made available for inclusion in meta-analysis.

## Authors Contributions

**Nancy E. Mayo** is the inventor of the Heel2Toe sensor. She designed the trial and developed the analysis plan, secured funding, oversaw the development of study material, and wrote up the results based on the analyses conducted by the data analyst, Lyne Nadeau.

**Kedar KV Mate** tested the Heel2Toe sensor as part of work done for his PhD. He participated in the development of study material, helped train assessors, delivered the intervention to selected English speaking participants, reviewed the results, provided edits to the manuscript.

**Lesley Fellows** provided input on the neuroscience underlying the mechanism of the Heel2Toe sensor.

**José Morais** provided lab space to conduct the study and participated in the testing of the Heel2Toe sensor and designing the material for the workbook.

**Madeleine Sharp** provided input on the neuroscience underlying the mechanism of the Heel2Toe and referred patients to the trial.

**Anne-Louise Lafontaine** provided input on the neuroscience underlying the mechanism of the Heel2Toe and referred patients to the trial.

**Edward Hill** is a co-inventor of Heel2Toe sensor, he built the sensors and designed the app for data capture and visualization. He made modifications to hardware and software as indicated.

**Helen Dawes** contributed expertise on the mechanism of action and on the interpretation of the results. She made comments on the manuscript.

**Ahmed-Abou Sharkh** developed the study material including the material for the RedCap database, trained assessors, co-ordinated the study, provided the intervention to participants, wrote up the results related to feasibility and usability as part of his work for a PhD. He reviewed the results and the manuscript.

## Data Availability

Pilot data only not available

## Acknowledgements

The study team wishes to thank the participants for their patience in testing a new technology. The team also acknowledges the contribution of Dr. Stanley Hum for the design and management of the RedCap database and Lyne Nadeau for conducting the analysis.

---

**Supplemental Table 1.** Average values for gait parameters during the baseline walking assessment as recorded from the Heel2Toe sensor for participants in each of the two groups.

|  | Heel2Toe Group (n=17) | Workbook Group (n=9) |
| --- | --- | --- |
| Number of recorded steps | 166.9 | 152.0 |
| Proportion “good” steps | 71.8% | 69.0% |
| **Heel Strike** |  |  |
| Angular velocity | -274.4 | -256.0 |
| Coefficient of variation | -52.8% | -56.5% |
| **Push-off** |  |  |
| Angular velocity | -257.6 | -279.3 |
| Coefficient of variation | -37.0% | -35.8% |
| **Power Phase** |  |  |
| Angular velocity | -1639.0 | -1902.02 |
| Coefficient of variation | -43.1% | -45.9% |
| **Foot Swing** |  |  |
| Angular velocity | 341.6 | 315.2 |
| Coefficient of variation | 20.9% | 19.9% |
| **Foot Swing Width** | 0.2813 | 0.3206 |
| Seconds | 0.2813 | 0.3206 |
| Coefficient of variation | 21.3% | 21.5% |
| **Balance Phase** |  |  |
| Angular velocity | 1789.5 | 2032.7 |
| Coefficient of variation | 29.8% | 27.7% |

A stride is 2 steps; Angular velocity is measured in degrees / second; Coefficient of variation is the ratio of standard deviation to the mean expressed in %

**Supplementary Table 2 – Part A:** Gait parameters at Baseline and 3 months for participants in Heel2Toe group with recorded data.

|  |  |  | Heel strike |  | Push off |  | Power phase AUC |  | Foot clearance |  | Swing time (sec) |  | Balance phase AAC |  |
| --- | --- | --- | --- | --- | --- | --- | --- | --- | --- | --- | --- | --- | --- | --- |
|  | Steps | % Good | AV | CV | AV | CV | AV | CV | AV | CV | AV | CV | AV | CV |
| Baseline | 160 | 71.9% | -239.7 | -52.8% | -187.9 | -32.3% | -1291.5 | -46.2% | 311.6 | 20.2% | 0.2774 | 19.9% | 1508.9 | 31.8% |
| 3 mon | 54 | 87.0% | -323.6 | -37.0% | -206.2 | -33.9% | -1651.8 | -32.3% | 389.6 | 14.1% | 0.2811 | 7.9% | 1897.5 | 14.8% |
| Baseline | 245 | 89.0% | -307.8 | -28.2% | -204.5 | -31.3% | -1381.9 | -43.0% | 308.0 | 18.3% | 0.2402 | 19.8% | 1451.6 | 30.9% |
| 3 mon | 114 | 88.6% | -294.5 | -32.7% | -276.6 | -21.9% | -1622.6 | -19.6% | 352.1 | 13.5% | 0.2735 | 9.5% | 1731.7 | 16.1% |
| Baseline | 55 | 47.3% | -146.7 | -87.5% | -211.7 | -42.9% | -884.5 | -61.2% | 277.8 | 28.7% | 0.2127 | 32.8% | 1132.3 | 47.5% |
| 3 mon | 243 | 84.8% | -320.0 | -41.8% | -229.6 | -42.4% | -1069.8 | -76.7% | 341.7 | 26.5% | 0.2193 | 33.4% | 1384.4 | 47.3% |
| Baseline | 56 | 62.5% | -328.3 | -74.7% | -385.0 | -37.2% | -1611.9 | -27.2% | 386.7 | 21.2% | 0.2446 | 18.1% | 1753.0 | 30.4% |
| 3 mon | 216 | 32.4% | -170.7 | -95.5% | -360.2 | -27.5% | -1259.3 | -36.5% | 449.2 | 16.3% | 0.2179 | 26.3% | 1830.8 | 31.0% |
| Baseline | 267 | 15.4% | -83.1 | -127.9% | -277.5 | -62.9% | -992.3 | -67.8% | 451.6 | 39.5% | 0.2399 | 35.6% | 1536.3 | 52.8% |
| 3 mon | 252 | 42.1% | -192.6 | -100.4% | -346.2 | -37.2% | -1482.5 | -43.7% | 593.2 | 26.0% | 0.2415 | 27.4% | 1944.5 | 41.3% |
| Baseline | 180 | 72.2% | -170.6 | -35.7% | -128.6 | -29.2% | -1075.2 | -18.1% | 240.0 | 22.3% | 0.2156 | 17.1% | 1078.3 | 18.5% |
| 3 mon | 17 | 100.0% | -268.1 | -18.0% | -170.7 | -11.9% | -936.6 | -21.6% | 285.8 | 22.0% | 0.2306 | 19.8% | 907.3 | 28.7% |
| Baseline | 307 | 94.8% | -428.8 | -32.0% | -220.5 | -43.3% | -1561.4 | -49.5% | 431.7 | 24.6% | 0.2336 | 17.5% | 1831.8 | 30.8% |
| 3 mon | 368 | 95.4% | -545.9 | -29.0% | -548.0 | -25.0% | -4224.9 | -21.0% | 493.2 | 16.5% | 0.2999 | 12.9% | 4584.7 | 19.7% |
| Baseline | 64 | 92.2% | -340.1 | -37.8% | -248.1 | -44.1% | -1308.4 | -138.9% | 463.9 | 25.3% | 0.2622 | 24.8% | 1766.0 | 31.6% |
| 3 mon | 348 | 94.8% | -357.7 | -32.2% | -555.1 | -8.0% | -3585.4 | 17.8% | 447.9 | 7.9% | 0.3206 | 7.9% | 3822.7 | 17.9% |
| Baseline | 23 | 78.3% | -320.0 | -61.7% | -208.2 | -80.2% | -1577.9 | -26.2% | 361.7 | 28.0% | 0.2574 | 11.3% | 1889.0 | 29.5% |
| 3 mon | 329 | 72.3% | -209.8 | -44.0% | -246.4 | -23.4% | -3407.2 | -33.2% | 228.2 | 8.5% | 0.3531 | 25.3% | 4122.8 | 18.4% |
| Baseline | 62 | 45.2% | -180.2 | -83.6% | -184.5 | -35.1% | -1336.7 | -33.4% | 304.9 | 23.2% | 0.2868 | 16.1% | 1479.5 | 29.2% |
| 3 mon | 321 | 76.0% | -308.0 | -52.3% | -261.2 | -27.7% | -1625.6 | -16.1% | 359.6 | 14.9% | 0.2899 | 10.2% | 1707.5 | 13.7% |
| Baseline | 240 | 87.5% | -409.3 | -43.5% | -270.8 | -43.8% | -1508.3 | -0.61588 | 472.6 | 28.8% | 0.2503 | 26.4% | 1708.4 | 43.9% |
| 3 mon | 323 | 90.1% | -274.6 | -29.7% | -341.6 | -15.7% | -4475.1 | -0.23049 | 260.3 | 10.6% | 0.3580 | 26.6% | 4723.3 | 17.4% |
| Baseline | 14 | 92.9% | -293.1 | -31.0% | -582.4 | -9.7% | -3631.4 | -24.0% | 371.9 | 11.4% | 0.3571 | 2.9% | 3487.7 | 14.2% |
| 3 mon | 13 | 53.8% | -156.7 | -30.5% | -310.9 | -25.1% | -3545.9 | -18.6% | 201.0 | 7.1% | 0.3538 | 5.6% | 3593.9 | 14.8% |
| Baseline | 258 | 30.2% | -118.6 | -50.7% | -217.1 | -15.6% | -3275.4 | -19.7% | 198.6 | 13.1% | 0.4104 | 28.9% | 3431.4 | 16.5% |
| 3 mon | 294 | 31.6% | -124.2 | -60.0% | -266.9 | -12.6% | -3522.0 | -21.4% | 206.8 | 10.2% | 0.4115 | 31.2% | 3940.9 | 14.0% |

Green: improvement by $\geq 10\%$ ; Yellow: no change; Orange: deterioration by $\geq 10\%$
Person-measures = 13 people \* 13 parameters = 169 person-measures

**Supplementary Table 2 – Part B:** Gait parameters at Baseline and 3 months for participants in Workbook group with recorded data.

|  |  | ngood |  | ntime | Heel strike |  | Push off |  | Power AUC |  | Foot Clearance |  | Swing width |  | Balance phase AAC |  |
| --- | --- | --- | --- | --- | --- | --- | --- | --- | --- | --- | --- | --- | --- | --- | --- | --- |
|  | Steps |  | % good |  | AV | CV | AV | CV | AV | CV | AV | CV | Seconds | CV | AV | CV |
| Baseline | 253 | 120 | 47.4% | 1.09 | -142.1 | -69.7% | -189.8 | -38.8% | -1345.8 | -46.5% | 317.0 | 24.4% | 0.2483 | 19.0% | 1435.4 | 28.6% |
| 3 mon | 266 | 91 | 34.2% | 1.18 | -139.9 | -76.5% | -200.1 | -37.0% | -1370.6 | -31.6% | 331.9 | 21.5% | 0.2502 | 13.8% | 1519.3 | 22.8% |
| Baseline | 55 | 50 | 90.9% | 0.82 | -483.5 | -35.6% | -295.7 | -46.3% | -1611.6 | -65.3% | 401.9 | 21.5% | 0.2338 | 23.1% | 1947.8 | 39.7% |
| 3 mon | 314 | 294 | 93.6% | 0.87 | -473.6 | -32.8% | -306.8 | -34.2% | -1792.9 | -53.8% | 445.0 | 22.3% | 0.2691 | 15.7% | 2161.2 | 27.1% |
| Baseline | 245 | 118 | 48.2% | 1.39 | -150.5 | -83.9% | -197.7 | -36.8% | -1176.1 | -25.2% | 309.5 | 15.4% | 0.2811 | 10.1% | 1502.1 | 15.9% |
| 3 mon | 249 | 59 | 23.7% | 1.36 | -123.8 | -73.1% | -193.0 | -33.9% | -1201.3 | -28.6% | 288.0 | 20.4% | 0.2561 | 11.2% | 1511.0 | 23.7% |
| Baseline | 19 | 15 | 78.9% | 0.92 | -252.8 | -48.9% | -229.7 | -23.3% | -1514.6 | -16.1% | 310.8 | 11.7% | 0.2726 | 11.0% | 1549.5 | 10.1% |
| 3 mon | 334 | 271 | 81.1% | 0.97 | -284.0 | -41.6% | -215.6 | -29.9% | -1439.0 | -29.0% | 263.1 | 18.4% | 0.2713 | 15.9% | 1540.4 | 23.7% |

Green: improvement by $\geq 10\%$ ; Yellow: no change; Orange: deterioration by $\geq 10\%$
Person-measures = 4 people \* 13 parameters = 52

## Supplementary Figures

**Supplemental Figure 1a.**
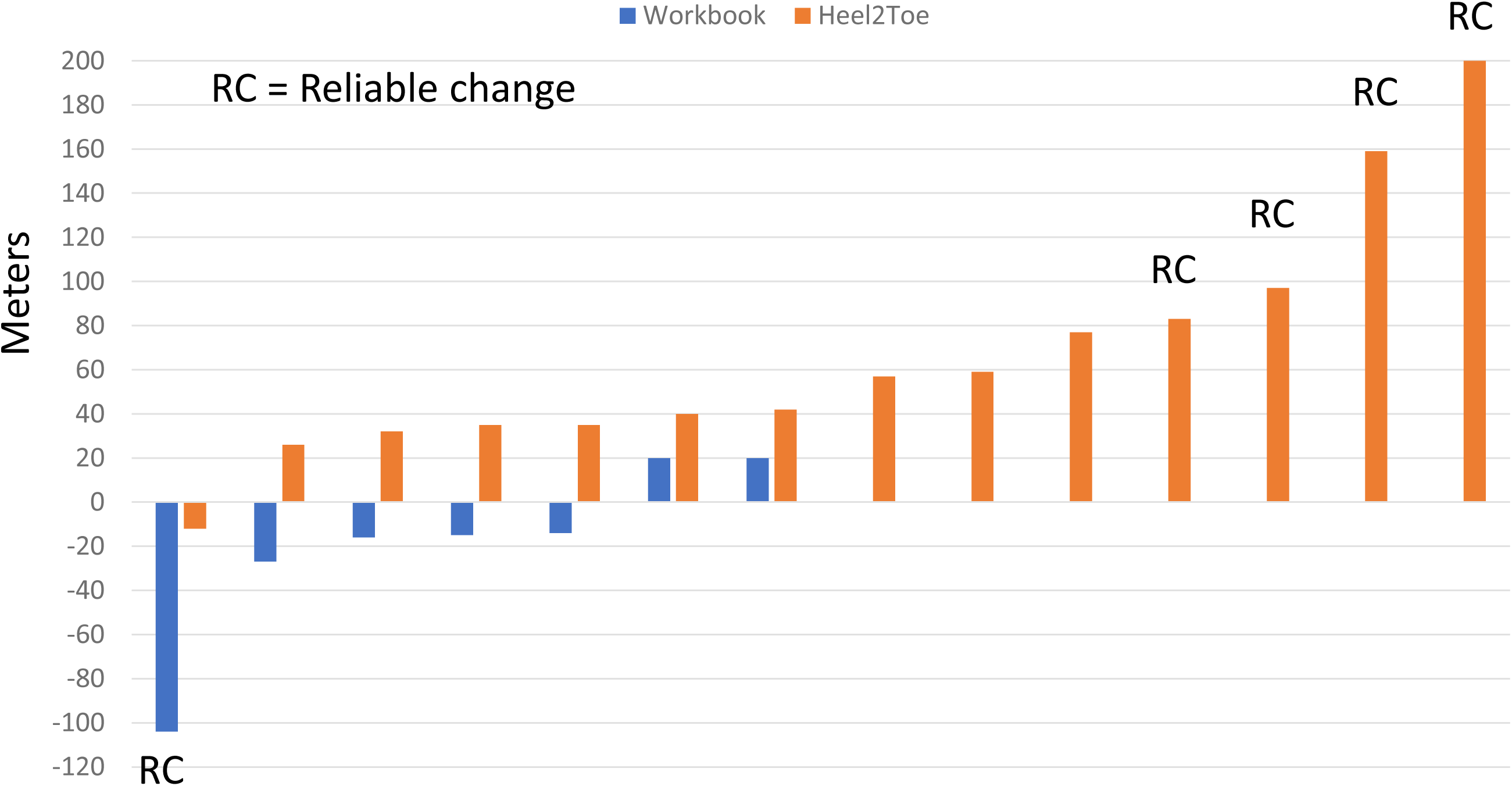
Individual values on change in 6MWT over the 3 month Intervention period for each group.

**Supplemental Figure 1b.**
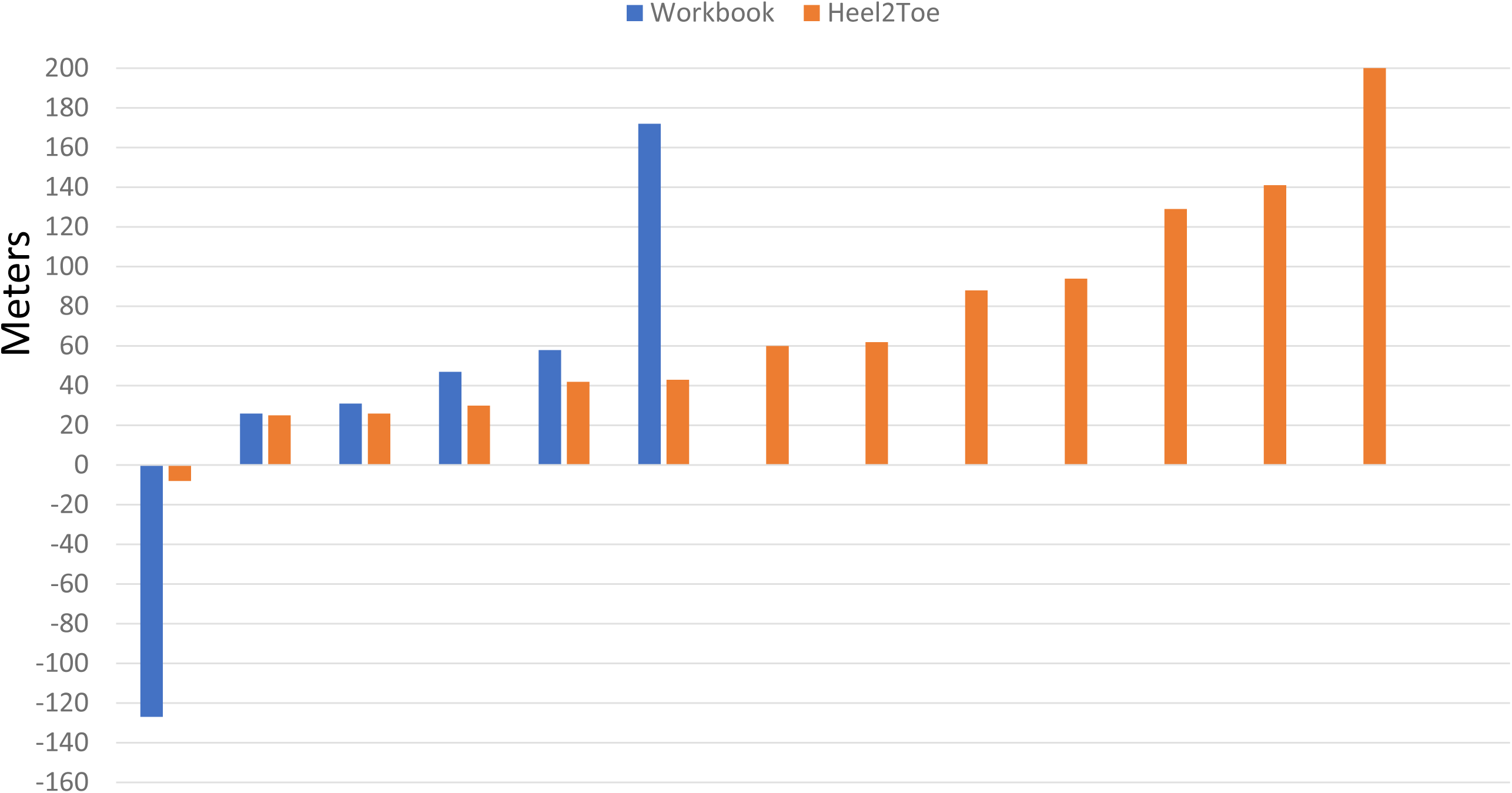
Individual values on change in 6MWT from baseline to 6 months for each group.

**Supplemental Figure 2.**
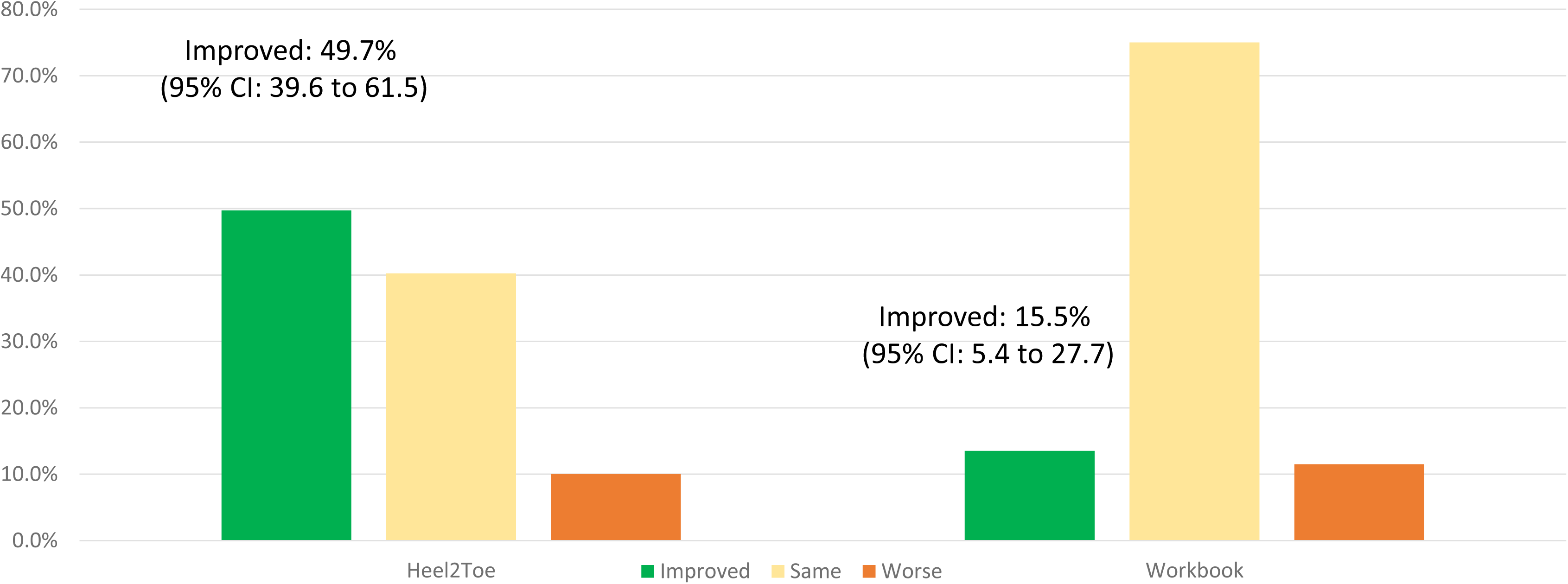
Proportion of person-gait measures showing improvement, no change, and deterioration over 3 months of active intervention for each group.

**Supplemental Figure 3.**
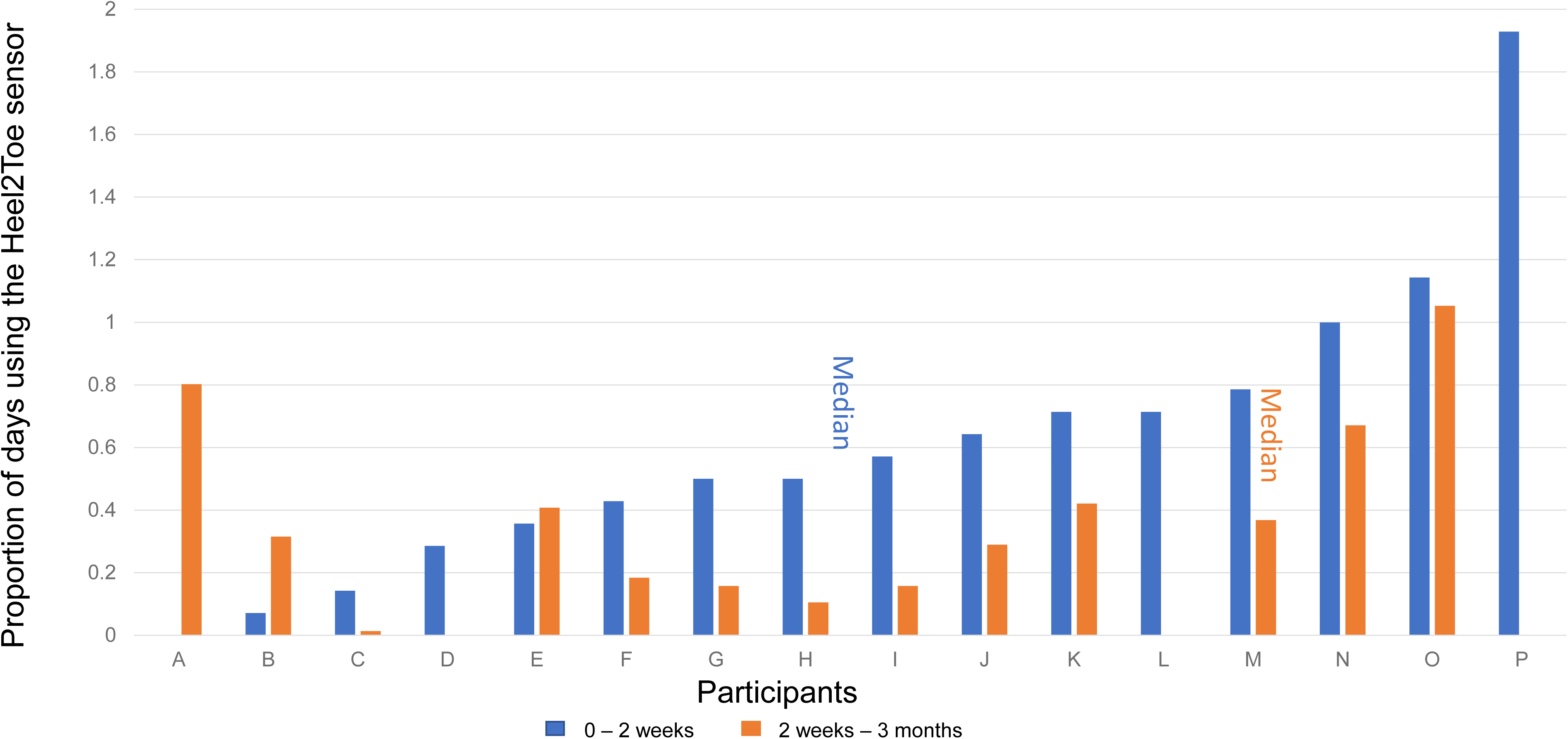
Distribution of Heel2Toe use for each participant over a 2-weeks training period followed by a 3-month home practice.

**Supplemental Figure 4.**
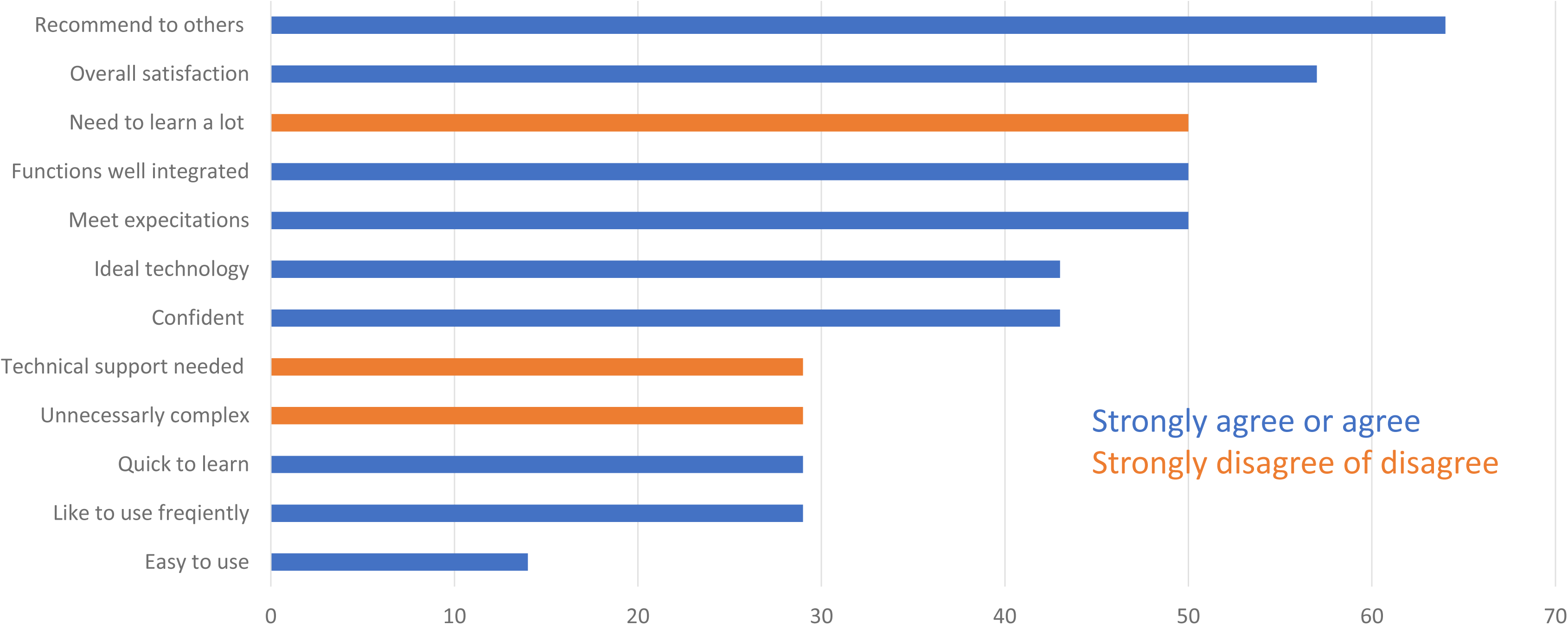
Proportion of the 14 participants using the H2T for the duration of the trial supporting its usability.

## References

1. Rangel-Barajas, C., I. Coronel, and B. Florán, Dopamine Receptors and Neurodegeneration. Aging and disease, 2015. 6(5): p. 349–368.

2. Wu, T., et al., Attention to Automatic Movements in Parkinson’s Disease: Modified Automatic Mode in the Striatum. Cereb Cortex, 2015. 25(10): p. 3330–42.

3. Gepshtein, S., et al., Dopamine function and the efficiency of human movement. J Cogn Neurosci, 2014. 26(3): p. 645–57.

4. Ayoubi, F., et al., Fear of falling and gait variability in older adults: a systematic review and meta-analysis. J.Am. Med Dir. Assoc., 2015. 16(1): p. 14–19.

5. Jankovic, J., Gait disorders. Neurol.Clin., 2015. 33(1): p. 249–268.

6. Ginis, P., et al., Focusing on heel strike improves toe clearance in people with Parkinson’s disease: an observational pilot study. Physiotherapy, 2017. 103(4): p. 485–490.

7. Lord, S., et al., Gait variability in Parkinson’s disease: an indicator of non-dopaminergic contributors to gait dysfunction? J Neurol, 2011. 258(4): p. 566–572.

8. Spildooren, J., et al., Turning problems and freezing of gait in Parkinson’s disease: a systematic review and meta-analysis. Disabil.Rehabil, 2018: p. 1–11.

9. Morris, M.E., Locomotor training in people with Parkinson disease. Phys Ther, 2006. 86(10): p. 1426–1435.

10. Alcock, L., et al., Step length determines minimum toe clearance in older adults and people with Parkinson’s disease. J Biomech., 2018. 71: p. 30–36.

11. Vadnerkar, A., et al., Classification of gait quality for biofeedback to improve heel-to-toe gait. Annu Int Conf IEEE Eng Med Biol Soc, 2014. 2014: p. 3626–9.

12. Vadnerkar, A., et al., Design and Validation of a Biofeedback Device to Improve Heel-to-Toe Gait in Seniors. IEEE J Biomed Health Inform, 2018. 22(1): p. 140–146.

13. Lovejoy, C.O., K.G. Heiple, and A.H. Burstein, The gait of Australopithecus. Am J Phys Anthropol, 1973. 38(3): p. 757–79.

14. Webber, J.T. and D.A. Raichlen, The role of plantigrady and heel-strike in the mechanics and energetics of human walking with implications for the evolution of the human foot. J Exp Biol, 2016. 219(Pt 23): p. 3729–3737.

15. Perry J.; Blumfield, J., Gait Analysis: Normal and Pathological Function 2010, New Jersey: Slack Incorporated.

16. McKay, M.J., et al., Spatiotemporal and plantar pressure patterns of 1000 healthy individuals aged 3-101 years. Gait Posture, 2017. 58: p. 78–87.

17. Mentiplay, B.F., et al., Lower limb angular velocity during walking at various speeds. Gait.Posture., 2018. 65: p. 190–196.

18. Rosati, G., et al., On the role of auditory feedback in robot-assisted movement training after stroke: review of the literature. Comput.Intell.Neurosci., 2013. 2013: p. 586138.

19. Kolb, B. and R. Gibb, Searching for the principles of brain plasticity and behavior. Cortex, 2014. 58: p. 251–260.

20. Cai, L., et al., Brain plasticity and motor practice in cognitive aging. Front Aging Neurosci., 2014. 6: p. 31.

21. Foerde, K. and D. Shohamy, The role of the basal ganglia in learning and memory: insight from Parkinson’s disease. Neurobiol.Learn.Mem., 2011. 96(4): p. 624–636.

22. Carvalho, L.P., et al., A new approach toward gait training in patients with Parkinson’s Disease. Gait Posture, 2020. 81: p. 14–20.

23. Mate, K.K., et al., Real-Time Auditory Feedback-Induced Adaptation to Walking Among Seniors Using the Heel2Toe Sensor: Proof-of-Concept Study. JMIR Rehabil Assist Technol, 2019. 6(2): p. e13889.

24. Berridge, K.C. and T.E. Robinson, What is the role of dopamine in reward: hedonic impact, reward learning, or incentive salience? Brain Res Brain Res Rev, 1998. 28(3): p. 309–69.

25. Eldridge, S.M., et al., Defining Feasibility and Pilot Studies in Preparation for Randomised Controlled Trials: Development of a Conceptual Framework. PLoS.One., 2016. 11(3): p. e0150205.

26. Eldridge, S.M., et al., CONSORT 2010 statement: extension to randomised pilot and feasibility trials. Pilot.Feasibility.Stud., 2016. 2: p. 64.

27. Goetz, C.G., et al., Movement Disorder Society-sponsored revision of the Unified Parkinson’s Disease Rating Scale (MDS-UPDRS): scale presentation and clinimetric testing results. Mov Disord., 2008. 23(15): p. 2129–2170.

28. Nasreddine, Z.S., et al., The Montreal Cognitive Assessment, MoCA: a brief screening tool for mild cognitive impairment. J Am.Geriatr.Soc, 2005. 53(4): p. 695-699.

29. Enright, P.L., et al., The 6-min walk test – A quick measure of functional status in elderly adults. Chest, 2003. 123(2): p. 387–398.

30. Taylor, M.J., Standardized Walking Obstacle Course: reliability and validity of a functional measurement tool. J Neurol Phys Ther, 1997. 21: p. 167.

31. Ng, S.S.M., et al., Reliability and concurrent validity of standardized walking obstacle course test in people with stroke. J Rehabil Med, 2017. 49(9): p. 705–714.

32. Rikli, R.E. and C.J. Jones, Functional Fitness Normative Scores for Community-Residing Older Adults, Ages 60-94. Journal of Aging and Physical Activity, 1999. 7(2): p. 162–181.

33. Starkstein, S.E., et al., Reliability, validity, and clinical correlates of apathy in Parkinson’s disease. J Neuropsychiatry Clin.Neurosci., 1992. 4(2): p. 134–139.

34. Rosenzveig, A., et al., Toward patient-centered care: a systematic review of how to ask questions that matter to patients. Medicine (Baltimore), 2014. 93(22): p. e120.

35. Cramp, F. and J. Byron-Daniel, Exercise for the management of cancer-related fatigue in adults. Cochrane Database Syst Rev, 2012. 11(11): p. Cd006145.

36. Jensen, M.P., C. Chen, and A.M. Brugger, Interpretation of visual analog scale ratings and change scores: a reanalysis of two clinical trials of postoperative pain. J Pain, 2003. 4(7): p. 407–14.

37. Kozlowski, A.J., et al., Evaluating Individual Change With the Quality of Life in Neurological Disorders (Neuro-QoL) Short Forms. Arch.Phys Med.Rehabil, 2016. 97(4): p. 650–654.

38. Franchignoni, F., A. Giordano, and G. Ferriero, Rasch analysis of the short form 8-item Parkinson’s Disease Questionnaire (PDQ-8). Qual.Life Res., 2008. 17(4): p. 541–548.

39. EuroQol, G., EQ-5D http://www.euroqol.org/ 2016.

40. Jayadevappa, R., R. Cook, and S. Chhatre, Minimal important difference to infer changes in health-related quality of life-a systematic review. J Clin Epidemiol, 2017. 89: p. 188–198.

41. Crimmins, E.M., et al., Changes in Biological Markers of Health: Older Americans in the 1990s. The Journals of Gerontology: Series A, 2005. 60(11): p. 1409–1413.

42. Verghese, J., et al., Quantitative gait markers and incident fall risk in older adults. J Gerontol A Biol Sci Med Sci, 2009. 64(8): p. 896–901.

43. Estrada, E., E. Ferrer, and A. Pardo, Statistics for Evaluating Pre-post Change: Relation Between Change in the Distribution Center and Change in the Individual Scores. Front Psychol, 2018. 9: p. 2696.

44. Sullivan, G.M. and R. Feinn, Using Effect Size-or Why the P Value Is Not Enough. Journal of graduate medical education, 2012. 4(3): p. 279–282.

45. Tate, J.J. and C.E. Milner, Real-time kinematic, temporospatial, and kinetic biofeedback during gait retraining in patients: a systematic review. Phys Ther, 2010. 90(8): p. 1123–34.

46. van Gelder, L.M.A., et al., The use of biofeedback for gait retraining: A mapping review. Clin Biomech (Bristol, Avon), 2018. 59: p. 159–166.

47. Mate, K.K.V., et al., Evidence for the Efficacy of Commercially Available Wearable Biofeedback Gait Devices: Consumer-Centered Review. JMIR Rehabil Assist Technol, 2023. 10: p. e40680.

